# Concordance between EMA Good Clinical Practice inspections and medical literature concerning drugs that have not received marketing authorization in the European Union: a meta-research survey

**DOI:** 10.1101/2025.05.09.25327294

**Authors:** Alexandre Terré, Ondine Becker, John P.A. Ioannidis, Florian Naudet

**Affiliations:** Department of Internal Medicine, Ambroise Paré Hospital, Boulogne-Billancourt, France; Sorbonne Université, Faculté de Médecine; Medical Oncology Department, European Hospital Georges Pompidou, Paris, France; Departments of Medicine, of Epidemiology and Population Health, and of Biomedical Data Science, and Meta-Research Innovation Center at Stanford (METRICS), Stanford University, CA 94305, USA; University of Rennes, CHU Rennes, INSERM, IRSET (Institut de Recherche en Santé, Environnement et Travail)-UMR_S 1085, CIC 1414 (Centre of Clinical Investigation of Rennes), Rennes, France; Institut Universitaire de France (IUF), Paris, France

**Keywords:** EMA, EPAR, Meta analyses, Post publication peer Review, Good Clinical Practice inspections

## Abstract

**Objectives:** To describe the differences between European Medicine Agency (EMA) Good Clinical Practice (GCP) inspection reports and medical literature on drugs with withdrawn or refused applications.

**Design:** A retrospective study comparing studies included in European Public Assessment Reports (EPARs) and the corresponding articles published in the medical literature.

**Data sources:** We screened all EPARs released by the EMA from inception to April 2024 for drugs that were refused or had a withdrawn application. In those EPARs, we looked for mentions of GCP inspections and details about them, then we searched for related publications on the inspected studies on bibliographic databases.

**Eligibility criteria:** All EPARs mentioning good clinical practice inspection were included in this survey.

**Data extracted:** Two reviewers independently gathered information on the GCP inspections and their findings, including the EMA’s opinion on their impact on the study data. The reviewers checked related publications for mentions of the inspection and any subsequent correction, retraction, or expressions of concern related to its findings.

**Main outcome measures:** The main outcome was the mention of the GCP inspection findings in the publication of the inspected studies. We also assessed whether there was any mention of these findings in a correction, retraction, or expression of concern.

**Results:** Out of 285 EPARs screened, 57 (20%) mentioned a GCP inspection. 58 distinct studies with inspections had 61 publications. For 17 publications the inspection occurred after the publication, for 20 the inspection happened before the publication, and for 24 the date of the inspection was unknown. Only 1 publication (2%) addressed the inspection findings. Moreover, there were no corrections, retractions, or expressions of concern related to inspection findings. Among the 61 publications, 26 (43%) were related with 24 distinct studies that had an inspection that casted doubts on data reliability, but none mentioned the inspections at or after the time of publication.

**Conclusions:** This meta-research survey indicates that health authorities’ GCP inspections are not reflected in the published literature, even when the inspections have put the data reliability in doubt. Journals should clearly specify which aspects of those studies are trustworthy and which ones are not.

**Trial registration:** osf.io/pa9fq/

## Introduction

Clinical trials are essential sources of information for healthcare professionals and public health policymakers. Due to the significant impact that their publication in medical literature can have on patient care, it is crucial that this information is trustworthy. Peer review prior to publication serves to identify and rectify errors, thereby ensuring the reliability of the literature. However, peer review has its limitations (1,2). It often faces limited resources, and external experts may struggle to comprehensively address all potential issues within the constrained time frame of reviewing a trial that may span several years. Sometimes criticisms or comments may escape this step, leading to the publication of clinical trials with errors (3–5). In these instances, the literature is expected to self-correct.

Health authorities such as the US Food and Drug Administration (FDA) and the European Medicines Agency (EMA) possess information that may assist in identifying and correcting errors in clinical trials before and/or after publication. They occasionally conduct Good Clinical Practice (GCP) inspections at clinical study sites, providing an in-depth evaluation of crucial steps of clinical trials, such as conduct, data collection, and analysis. The detail provided by such investigations provides material that exceeds what medical journals can offer with peer-review. The report of these inspections, when identifying deficiencies, is crucial and should be known by the end-users. However, it seems that these inspection reports are disconnected from the evidence published in the medical literature. In 2015, Seife identified 57 clinical trials resulting in 78 publications where a GCP inspection by the FDA found deviations from GCP recommendations that could cast doubt on the quality of the information presented in the corresponding publications (6). Yet, in 2015, at the time of Seife’s publication, 96% of the articles he identified did not mention these inspections either themselves or through an expression of concern or erratum (6).

This question has not been examined through clinical trials assessed by the EMA. Unlike the FDA, the EMA does not directly publish GCP inspection reports. However, these are referenced in the European Public Assessment Reports (EPAR) issued by the Committee for Medicinal Products for Human Use (CHMP), especially when the outcome of a GCP inspection influences the CHMP’s decision to approve or deny a marketing authorization (7). The objective of our study was to determine if the EPARs for drugs submitted to the EMA contain information from GCP inspections that contradicts the medical literature data. To increase the chances of identifying GCP inspections that raised concerns, we focused on marketing applications that were either refused or withdrawn.

## Methods

The methods of this descriptive study were specified in advance in a protocol registered on the Open Science Framework on 31^st^ July 2024 (https://osf.io/pa9fq/). This report provides details and reasons for any deviations from the initial plan. As no reporting guideline exists for such a meta-research survey, we relied on PRISMA (8) which we considered as the most appropriate reporting guideline.

### Eligibility criteria and information sources

We included EPARs published from the 17th of July 2000 (date of publication of the first EPAR) up to the 1^st^ of April 2024, concerning drugs refused or for which the marketing application has been withdrawn. Among those EPARs, any study with a GCP inspection was included. The EPARs are labelled under the commercial name of the drug on the EMA website, thus here we use this name in our article and for EPARs citations.

### Search strategy

The assessments of the CHMP are publicly available on the EMA website, under the name of European public assessment reports (EPAR) (9). Using the search function on the EMA website, we searched the published assessments reports using the filters “human,” “Medicines,” “Withdrawn application,” “refused.”

For each identified assessment, we searched the available documents on the EMA website for mentions of a GCP inspection conducted by the EMA by searching the terms “Good clinical practice” or “GCP” used in these reports. We preferably used the document “EPAR - Public assessment report.” We also searched, when available, in the paragraph: “Steps taken for the assessment of the product.” In the absence of this document, the other documents presented on the EMA website (“withdrawal letter”, or “Question-Answer”) were also screened.

### Selection and data collection processes

Screening of EPARs and data collection of verbatim quotes were done independently by two authors (AT and OB). Contrary to our initial protocol, discrepancies were first resolved through consensus before being arbitrated by a third author (FN).

### Data items

#### Assessment report data

For each identified EPAR, we recorded the opinion date, the commercial name of the drug, the international non-proprietary name, whether the application was refused or withdrawn, and whether a GCP inspection conducted by the EMA was mentioned.

When a GCP inspection was identified, we extracted whether it was described and if deviations were mentioned in the EPAR. For each study subject to a GCP we noted if the results of the inspection impacted the efficacy criterion, the safety/tolerance profile of the drug, or an ethical rule. The deviations observed by the EMA are classified as “critical,” “major,” or “minor”. Critical findings are defined as “conditions, practices, or processes that adversely affect the rights, safety or wellbeing of the subjects and/or the quality and integrity of data”. Major findings are defined as “conditions practices or processes that might adversely affect the rights, safety or wellbeing of the subjects and/or the quality and integrity of data. Major observation are serious deficiencies and are direct violation of GCP principles”. Minor deviations are defined as “conditions, practices or processes that would not be expected to adversely affect the right, safety or wellbeing of the subjects and/or the quality and integrity of data” (10).

We extracted information following this classification by using the exact terms employed in the report. As the EPARs sometimes describes the number of deviations observed by category, we extracted the number of “critical” deviations per GCP inspection. If the EPAR detailed the verbatim elements of the GCP inspection influencing its decision, these elements were collected in free text. Finally, we looked at the conclusions issued by the CHMP concerning the impact of deviations eventually observed on the results presented. To clarify this last aspect, we had to refine data analysis process during our study because the classification of the findings as “critical”, “major” or “minor” was frequently missing or was not related to the reliability of the data as assess by the CHMP. Using the verbatim text presented in the EPAR we extracted the opinion of the EMA on the effect of the GCP findings on the reliability of the data. The findings were classified as “impacting the reliability of the data”, or “not impacting the reliability of the data”. If the opinion was not described, or ambiguous, the opinion was classified as “unknown”. We also examined whether the GCP inspection referenced any deviations from ethical guidelines.

#### Subsequent publications

For each mention of a GCP inspection, we searched if one or more of the corresponding studies were published in a peer-reviewed medical journal. The EPARs were searched under “Clinical efficacy” and “Main studies” for any EU Clinical Trials Register or ClinicalTrials.gov trial identification number or study number used by the sponsor. If only a sponsor’s study number was available, we searched in Google for its match with a Clinical trial number, using the sponsor’s study number and international non-proprietary name of the drug. Then, we searched in Google Scholar and study registries for the clinical studies published in medical journals using the identification number. We considered both studies published as articles and as abstracts or conference presentations and recorded separately the results published in articles and those posted in abstracts or conference presentations. In the absence of an identification number, we searched in free text the descriptive elements of the study using the drug’s name (commercial or international non-proprietary name), study design regarding randomization, comparator, the presence of blinding (single or double), and the medical indication (disease name with stage if specified) in the Google search engine. We used the number of patients and the inclusion dates to confirm that the studies found correspond to those mentioned in the EPAR. We collected all retrieved publications even when there were multiple publications for a given trial. For each identified publication, we examined references to the GCP inspection and its conclusions. We also searched for corrections, retractions or expression of concerns mentioning the GCP findings on Pubmed and on the journal website. This outcome was added during the research because many articles were published before the EPAR, so the initial version of the article was not expected to mention the GCP inspection. During the study, a search was added on the Pubpeer website (11) for any comments posted about the articles.

#### Dissemination in meta-analyses, reviews, and citation count

For each identified publication, we used the Web of Science search engine to collect the citation count of each study and specifically study the number of meta-analyses citing them. This was done between the 2nd of January and the 5th of February 2025.

#### Detailed inspection reports

To obtain comprehensive details of each GCP inspection revealing critical or major deviations and resulting in a scientific publication, we requested the complete GCP inspection report from the EMA. This request was made in accordance with Article 7 of Regulation (EC) No 1049/2001 regarding public access to European Parliament, Council, and Commission documents (the Regulation) and Section 3 of the Annex to the “European Medicines Agency policy on access to documents - POLICY/0043”(12). According to this policy, all documents from EU Institutions and European decentralized Bodies such as the European Agencies are accessible to the public. For each request we noted the date on which the request was made. We noted how long it took the EMA to respond to our request, and the answer we received. We planned that if there was no response after one year, we would consider the EMA did not address our request.

### Outcomes

The primary outcome was the mention of the deviation and/or of the EMA GCP inspection in the publication. This definition had to be enhanced by incorporating information on corrections, retractions, or expressions of concern related to the GCP findings. The secondary outcomes were 1/ description of the deviations in the EPAR, both qualitatively and quantitatively, 2/ discordance between the deviations described in the EPAR and the publication, 3/ description of publications in terms of journal and date, 4/ dissemination of the studies with data reliability issues identified by the EMA in journals and meta-analyses and 5/ comments on Pubpeer about the GCP inspection (this outcome was added during the study).

### Analysis

We described quantitative variables using means (± standard deviations, SD) or medians (interquatrile range, IQR), as appropriate. We described qualitative variables using numbers and percentages. Verbatim text is presented to describe findings of the GCP inspections.

### Patient involvement

Patients were not involved in formulating the research question or the outcome measures, nor did they participate in the design and implementation of the study. The findings of this project will be presented at the first meeting of the external committee for RestoRes (Research integrity in biomedical research), funded by the French Agence Nationale de la Recherche under grant agreement ANR-23-CE36-0006. This committee, which includes patient representatives and citizens, will be consulted to discuss the implications of the findings and determine the concrete actions to be implemented following this research project.

### Open access

Data and code to reproduce the analysis are available on the Open Science Framework (https://osf.io/pa9fq/files/osfstorage). EPARs used for the study are cited via the EMA link.

## Results

### Selection of EPARs, GCPs, published articles

Between 17th of July 2000 (date of publication of the first EPAR) up to April 2024 we screened 285 EPARs of refused or withdrawn applications. Fifty-seven (20%) of those were included (13–69) (presented in the supplementary references section) as they mentioned a GCP inspection. Those 57 EPARs referred to 74 different studies, 54 (73%) with a registered identification number (NCT or EudraCT). Among the 74 studies, 58 (in 45/57 EPARs) could be related to at least one publication. Among those, 52/58 studies (90%) were related to only one publication, and 6/58 (10%) to 2. Three publications reported results of 2 studies. This resulted in a total of 61 publications (70–130). One drug, masitinib, was related to 3 different EPARs (39,54,58), all mentioning critical findings in 3 studies, all casting doubts on the data integrity, and leading to 4 publications (70,94,101,102). **Figure 1** describes the selection process.

**Figure 1:**
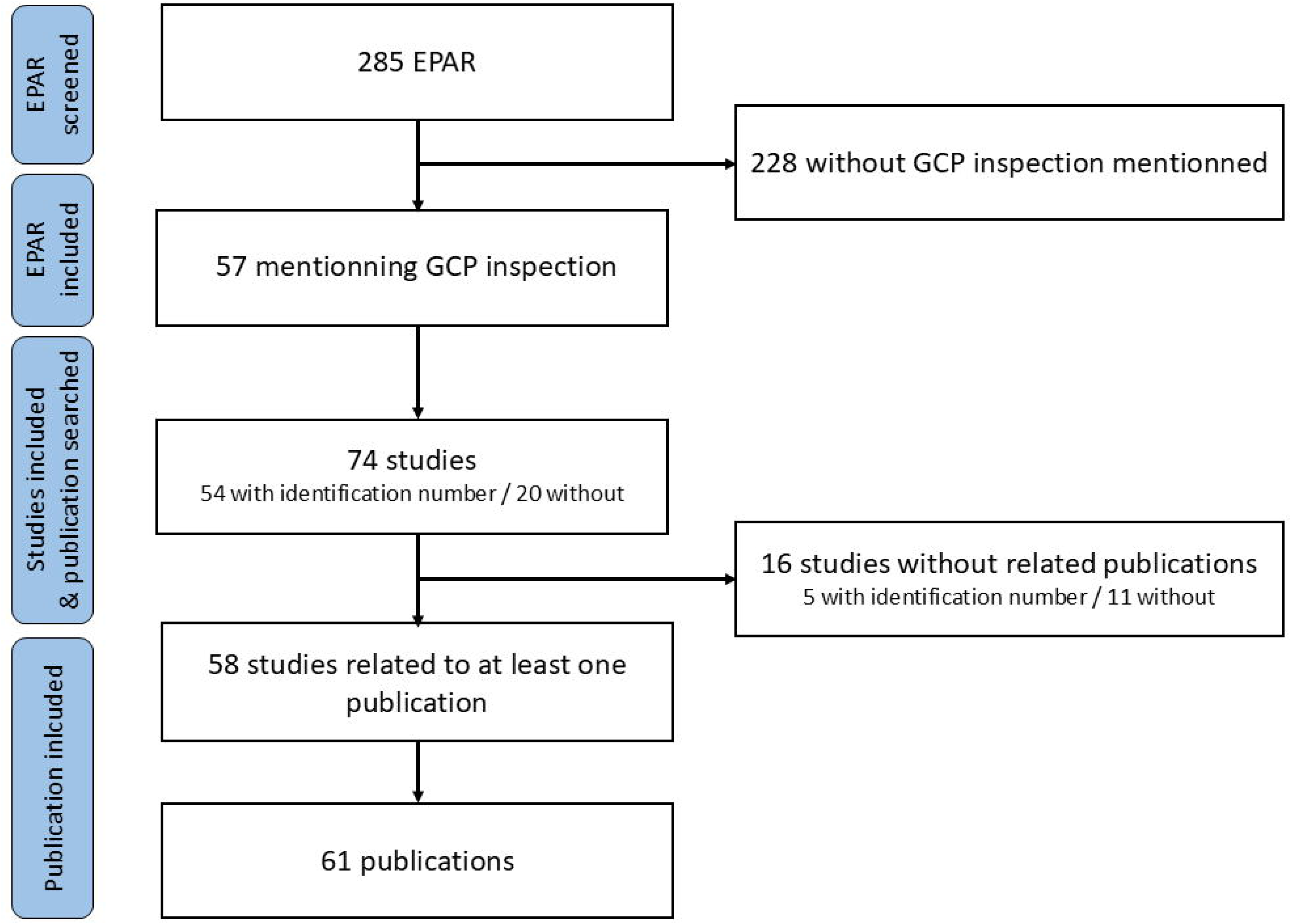
Flow chart of the study.

### Description of the deviations in the EPARs

26/57 EPARs (46%) did not specify whether the GCP inspection findings were classified as critical, major, or minor. 20/57 (35%) EPARs reported “critical” findings during the GCP inspections, and 27 (47%) reported “major” findings (presence of critical and major findings are not exclusive within an EPAR, based on EMA definitions). As described in 34/57 (60%) EPARs, the deviation affected the efficacy outcome in 26/34 (76%), the safety in 20/34 (59%) and ethics in 4/34 (12%) (deviations can be observed in multiple aspects of the study within an EPAR). Among the 57 EPARs, the GCPs findings were classified as “impacting the reliability of the data” (in at least on study mentioned in the EPAR) for 28/57 (49%) and “not impacting the reliability of the data” for 19/57 (33%). For 10/57 (18%), details were insufficient to judge the impact on reliability of the data from included studies. **Figure 2** describes the correspondence between EMA’s classification and the impact on data reliability. Information on the number of deviations per inspection was missing in 39/57 (68%) EPARs. A median of 0 (IQR: 0-2.75) critical deviations were identified in the remaining reports. In the EPARs mentioning critical deviations (n=20/57; 35%)), the number of critical deviations was described in 6/20 (30%). A median of 5 (IQR 3.25-9.75) critical deviations were reported. Those results are summarized in **Supplementary Table 1**.

**Figure 2:**
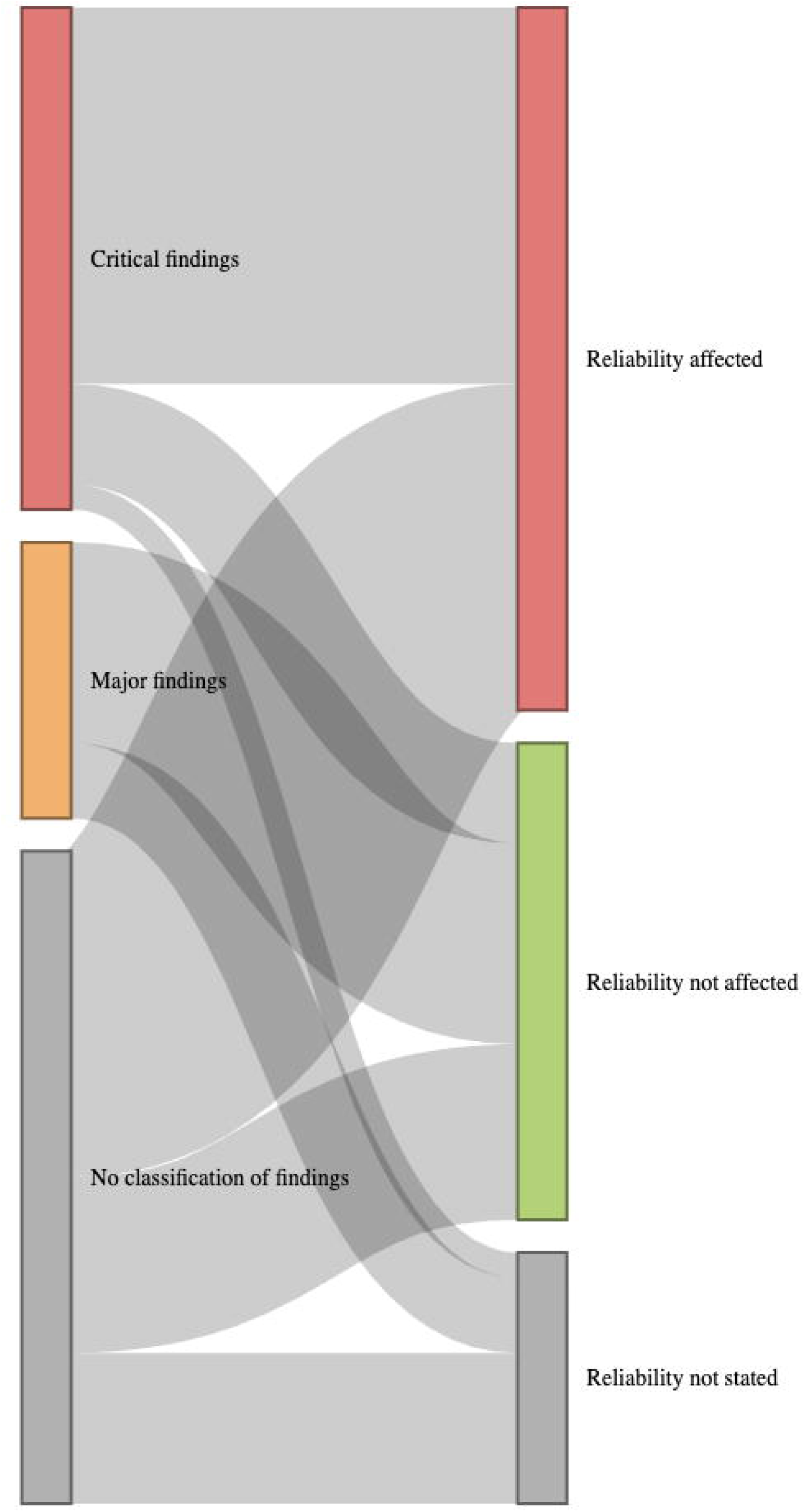
Sankey diagram showing the classification of Good Clinical Practice inspection in relation to reliability assessment by the Committee for Medicinal Products for Human Use. The data and code necessary to replicate the figure can be accessed through the Open Science Framework (https://osf.io/pa9fq/).

### Mention of the deviation and/or of the EMA GCP inspection in the published record

The information on all publications is available in **Supplementary Table 2**. Regarding our primary outcome, only 1/61 (2%) mentioned GCP inspection findings in the publication (131). This article discussed naproxcinod for knee osteoarthritis. The EPAR excluded data from a site due to GCP breaches, including lack of informed consent and recruitment of site personnel or their associates. It noted in the publication: “1 site excluded due to quality issues, and 1 patient excluded having been randomized at 2 sites.” (131). The number of patients in the EPAR and the article matched (28). However, as detailed in **Figure 3**, the corresponding EPARs were released in the public sphere after the publication 45/61 (74 %) publications. Among those, the GCP inspection occurred after publication for 17/45 (38%) and occurred before the publication in 8/45 (18%). For the remaining 20/45 (44%) the date of the GCP inspection was unknown. In such cases, we found no corrections, expressions of concern, retractions, related to the GCP inspection findings. One of those studies (102) had a correction related to an undisclosed conflict of interest (132).

**Figure 3:**
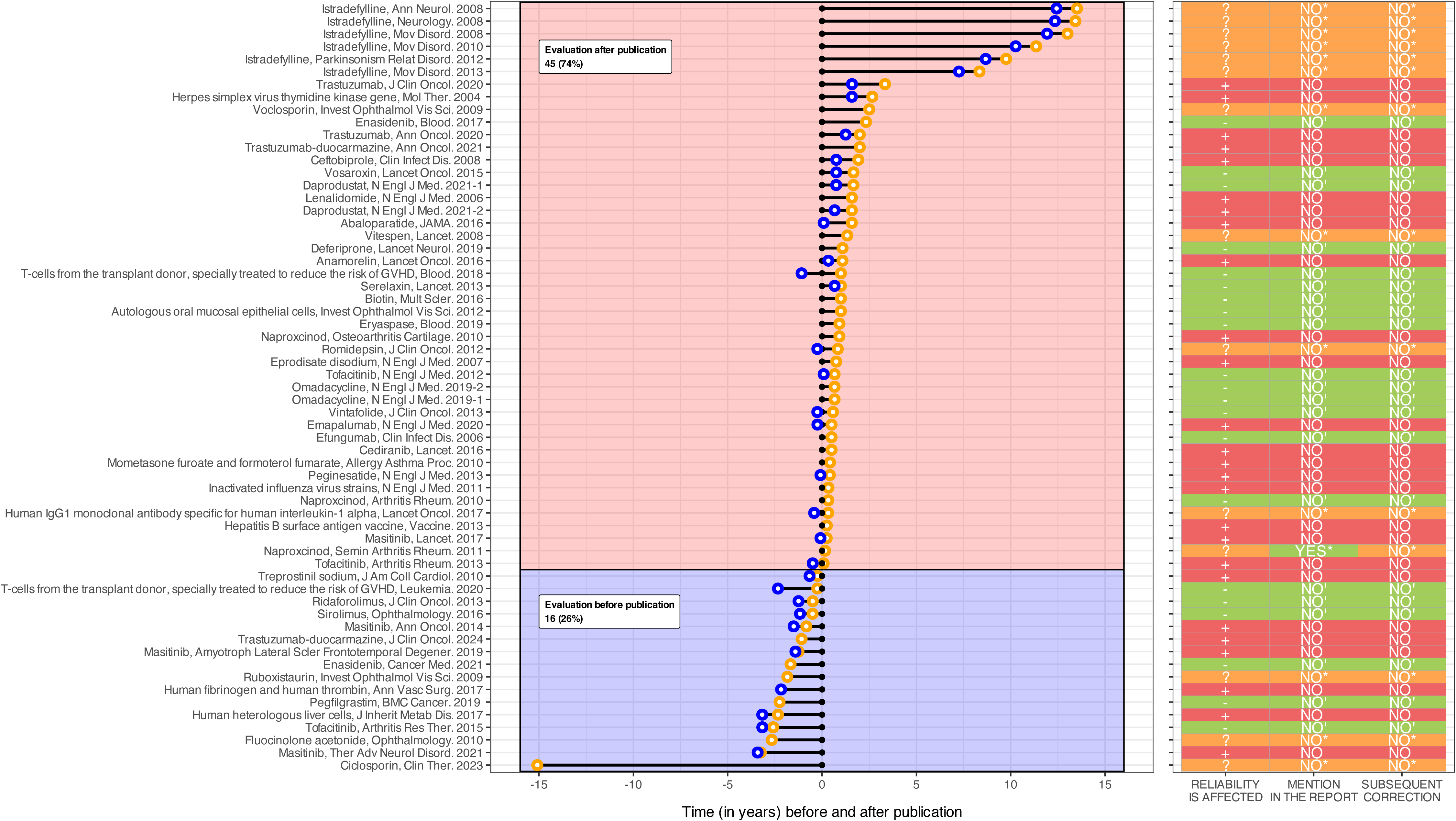
Chart showing the date of the inspection report (blue points), the date of the publication of the European public assessment report (orange points), and the date of the publication in a medical journal (black points), along with EMA’s evaluation regarding the reliability of the findings, mention in the publication, and subsequent correction, retraction, or expression of concern. − = data reliability non affected; + = data reliability affected; ? = unknown impact of the inspection’s findings on data reliability; ‘ = no mention of the findings expected; * = Findings that may or may not be relevant to report, as it was unclear whether the reliability was affected. The data and code necessary to replicate the figure can be accessed through the Open Science Framework (https://osf.io/pa9fq/).

For 21 publications (36%), the EPAR indicated that the GCP inspection did not affect the reliability of the data. For 14 publications (20%), the impact on data reliability was not described or was ambiguous. For 26 publications (43%) the EPAR indicated that GCP findings raised concerns about data reliability. The results are summarized in **Figure 3**. Only 2/61 (3%) publications (94,102) had comments on Pubpeer concerning the CGP inspection. They both were related to studies with critical deviations and data reliability issues (54,58). Actualy, both of those comments were posted by a member of our team before the start of our meta-research study.

Among the 16/74 (22%) unpublished studies, the EPAR indicated that the GCP inspection did not affect the reliability of the data in 5/16 (31%). For 5/16 (31%), the impact on data reliability was not described or was ambiguous. For the remaining 6/16 (38%), the EPAR indicated that GCP findings raised concerns about data reliability (**Supplementary Table 3**).

### Deviations, publications and dissemination of studies with data reliability issues

For the 26 studies with data reliability issues, the verbatim text of the EPAR mentioning the main findings and the journals in which those studies were published are presented in **Table 1**. The only journals with multiple published articles of this category were *The New England Journal of medicine* (n=6, 23%), *The Lancet* (n=2, 8%), *Annals of Oncology* (n=2, 8%). Those studies with reliability issues were published between November 2004 and October 2024 and had a median citation number of 135 (IQR 46;308.5) and a median citation number by meta-analyses of 5 (IQR 1;9.5).

**Table 1:**
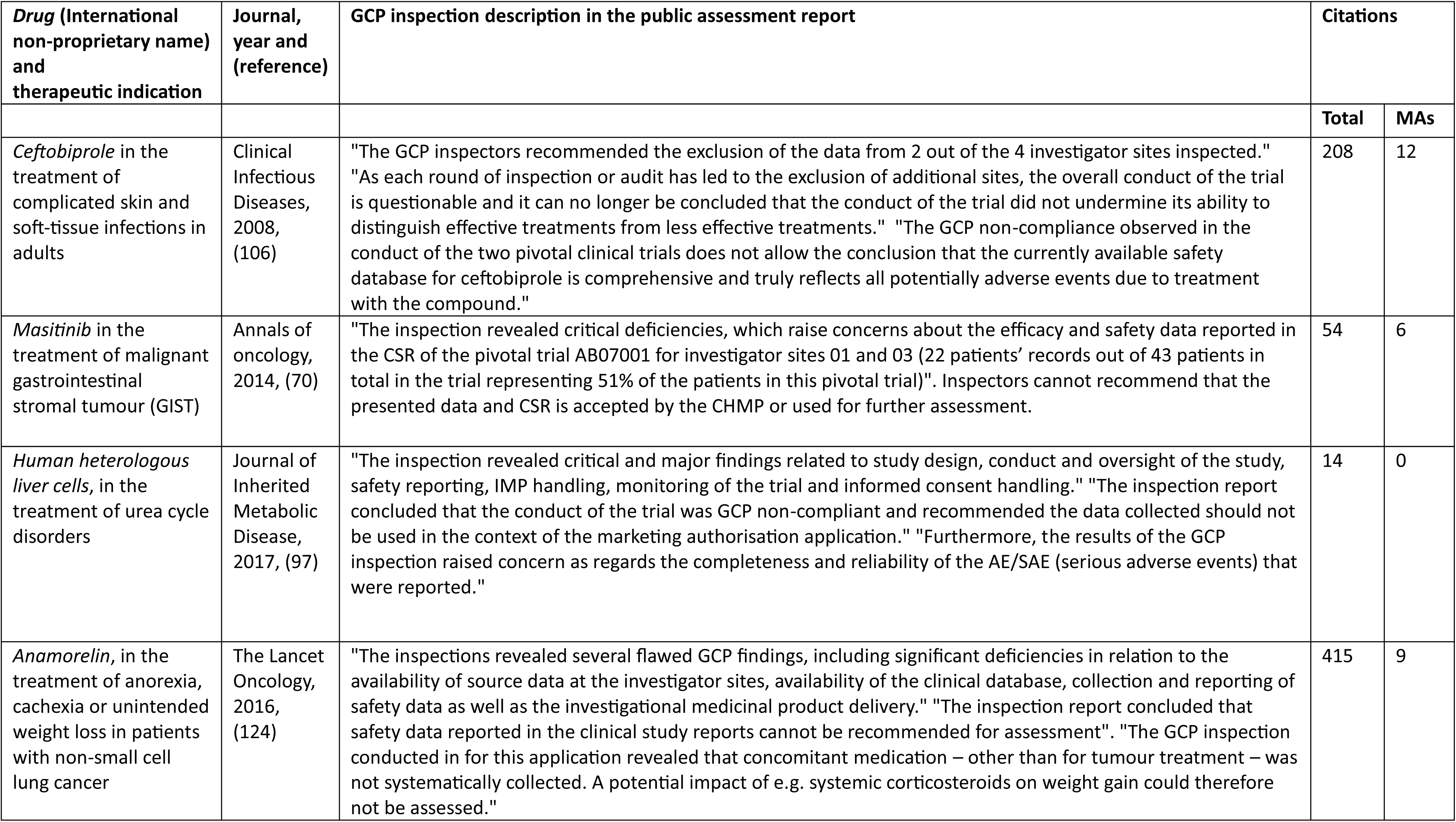
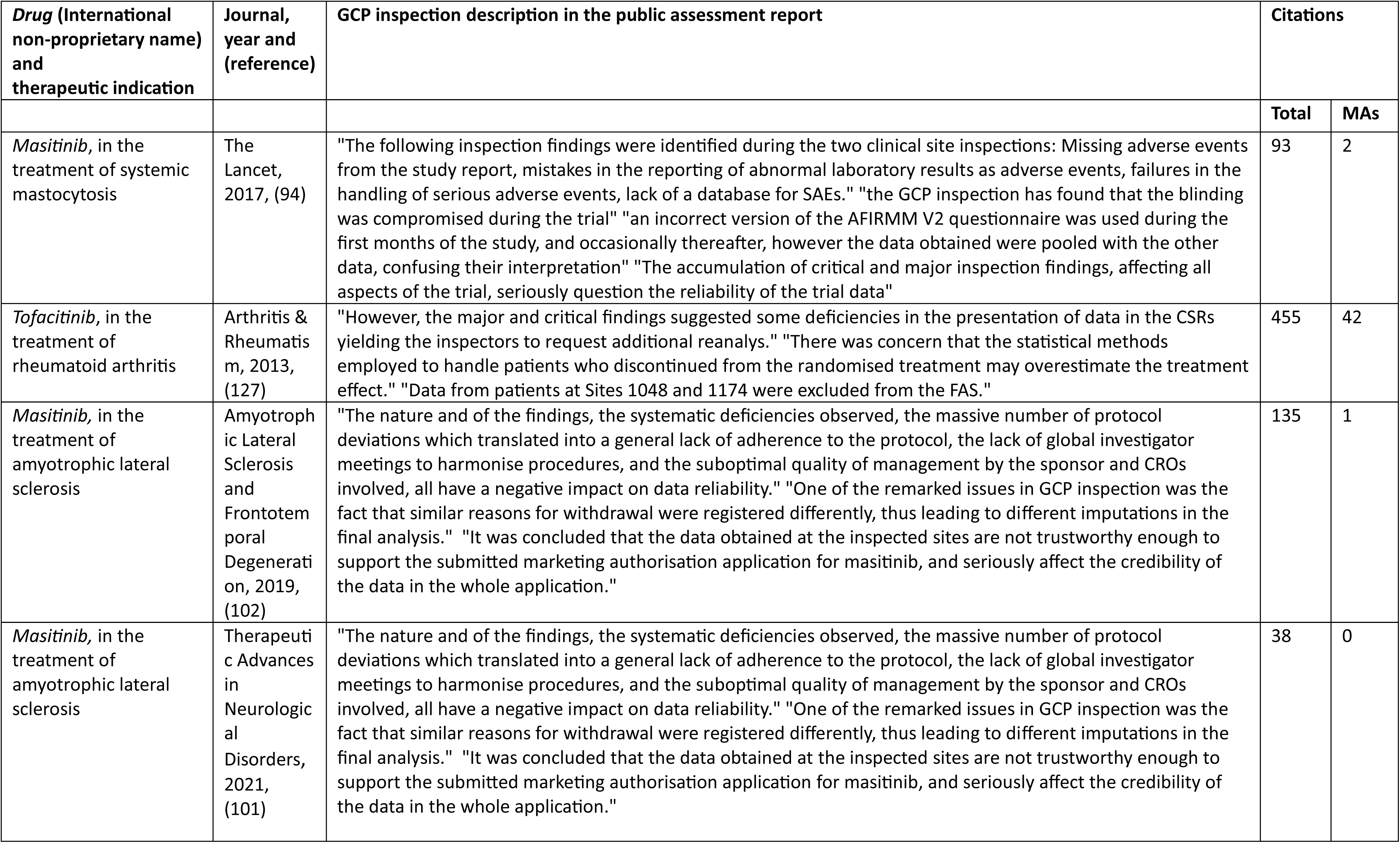
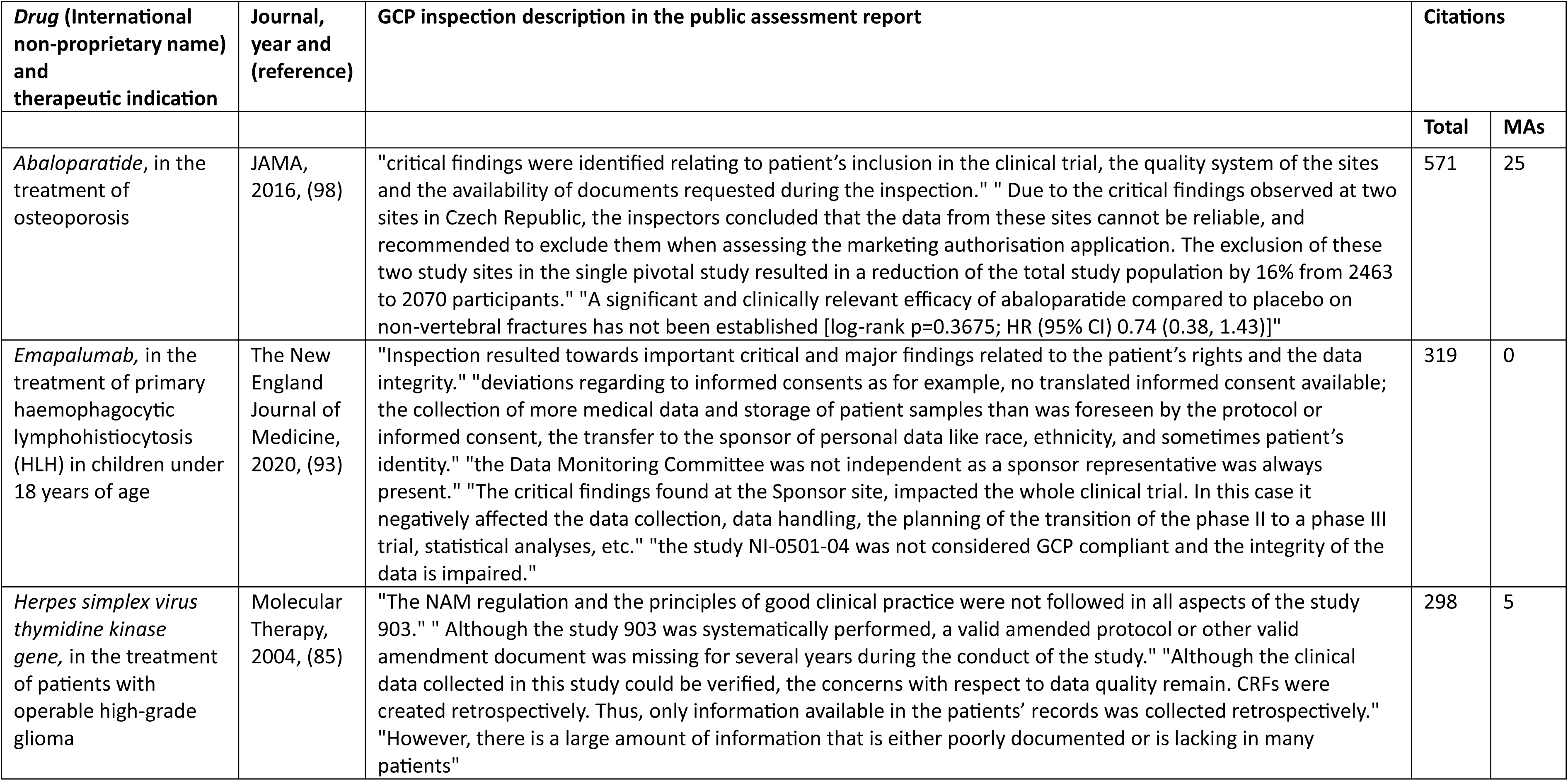
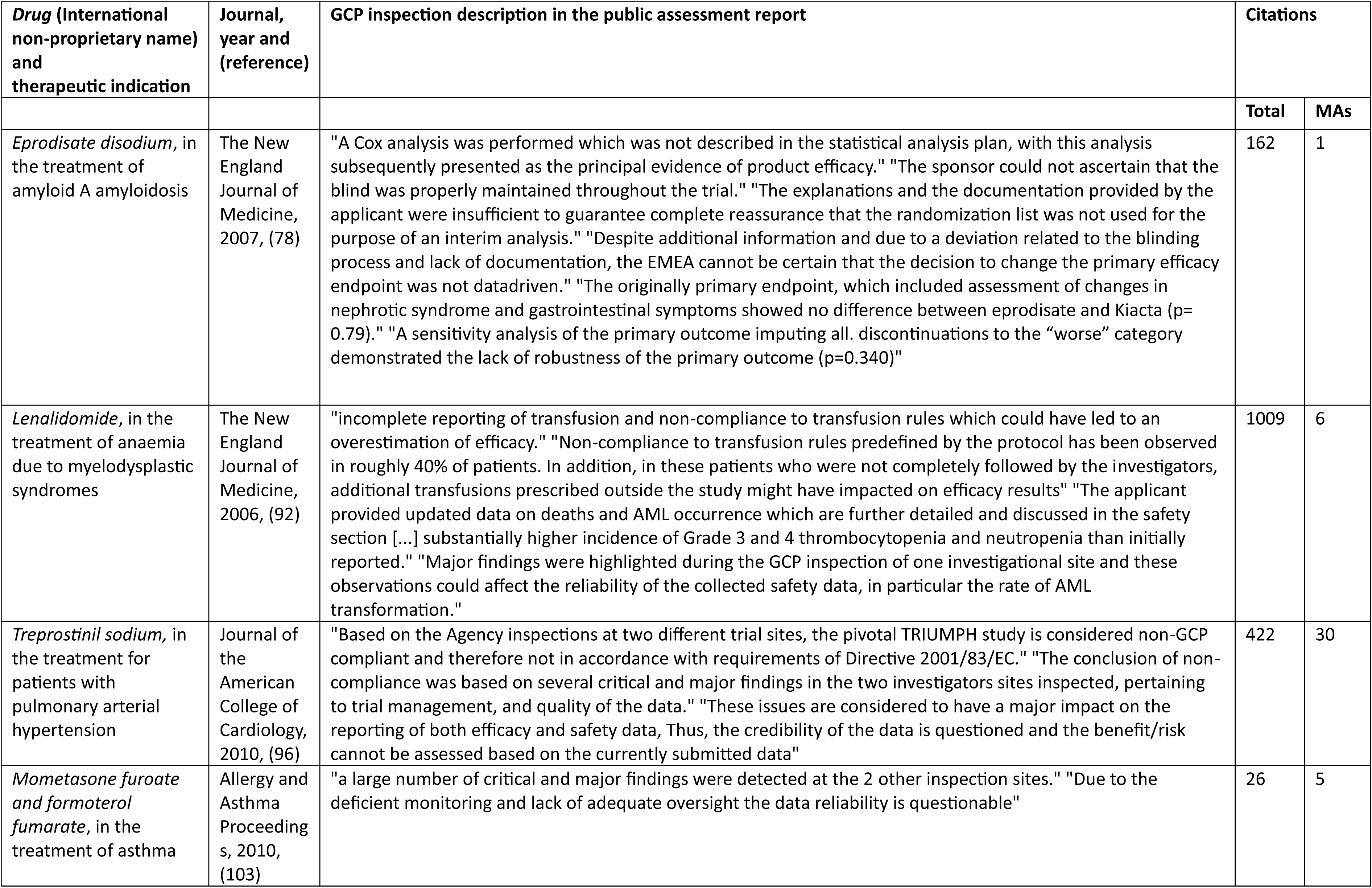
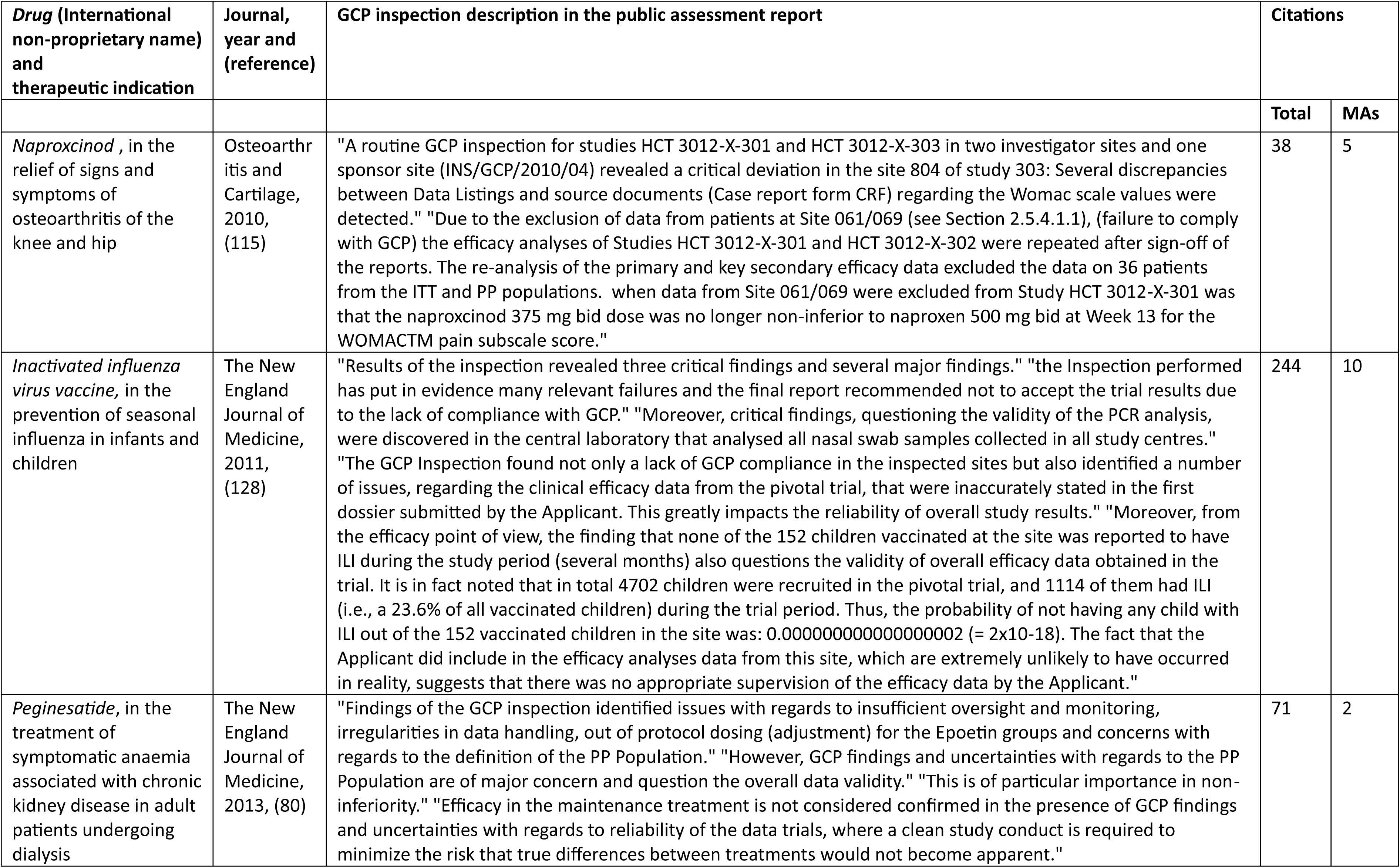
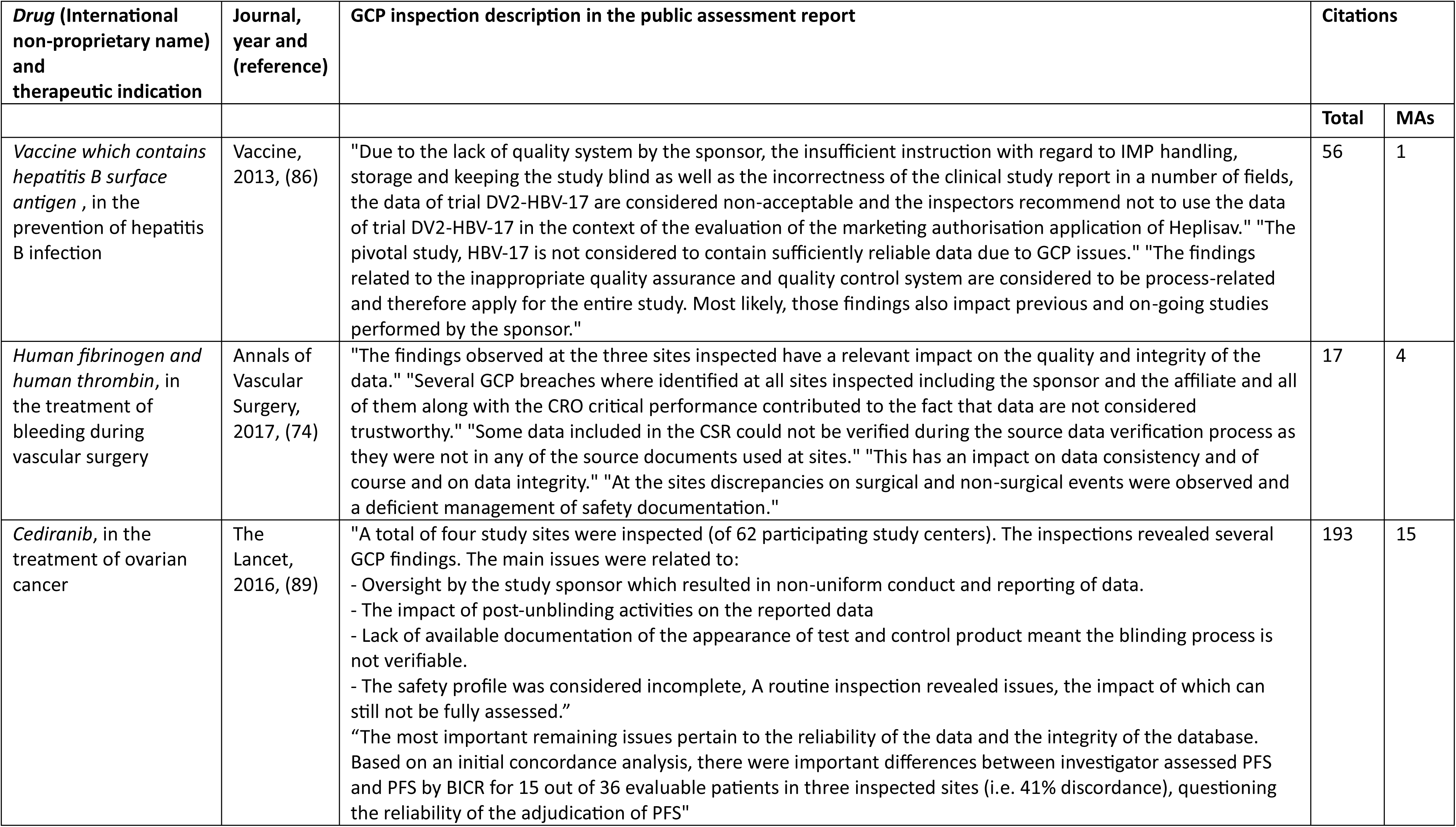
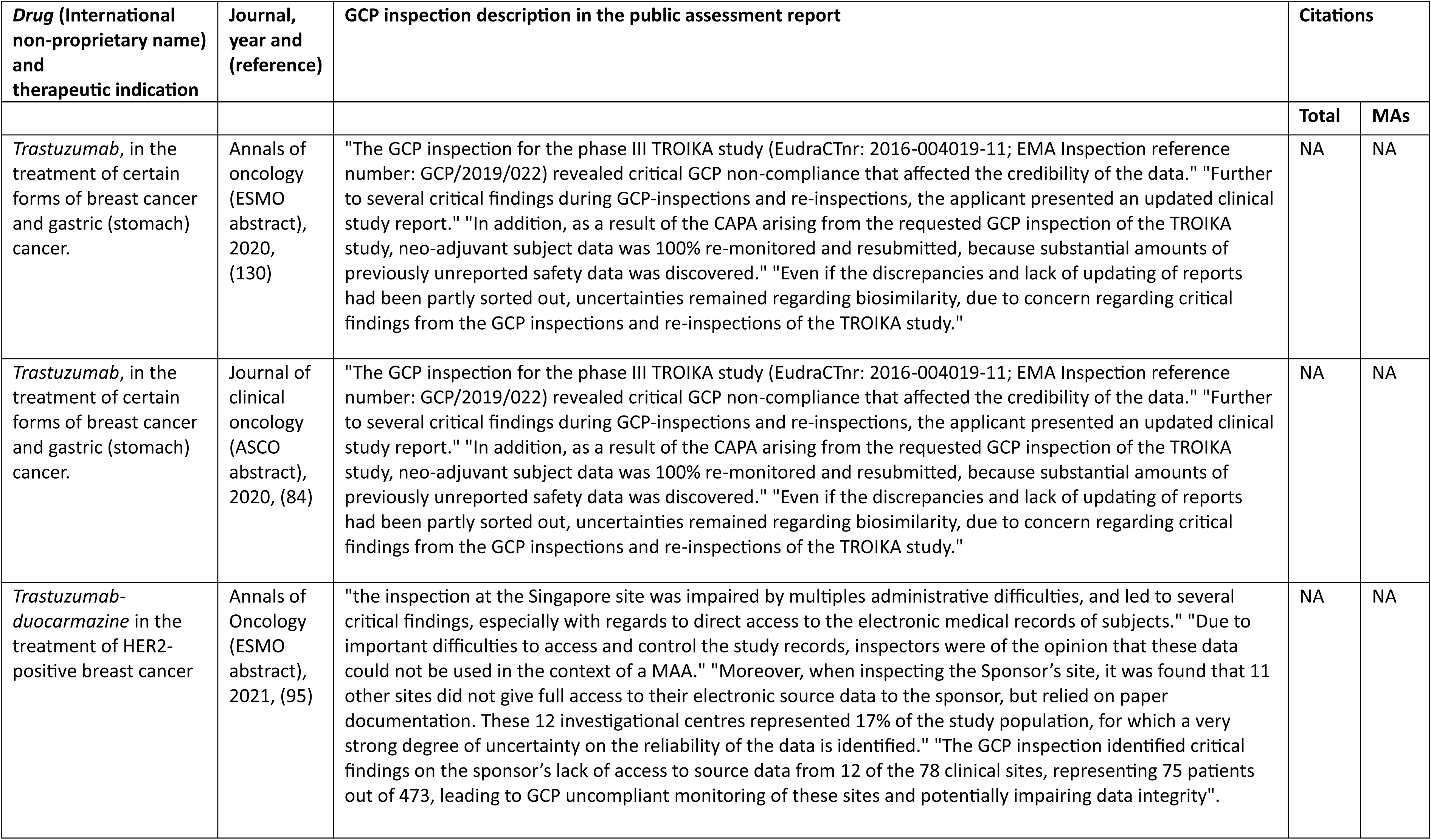
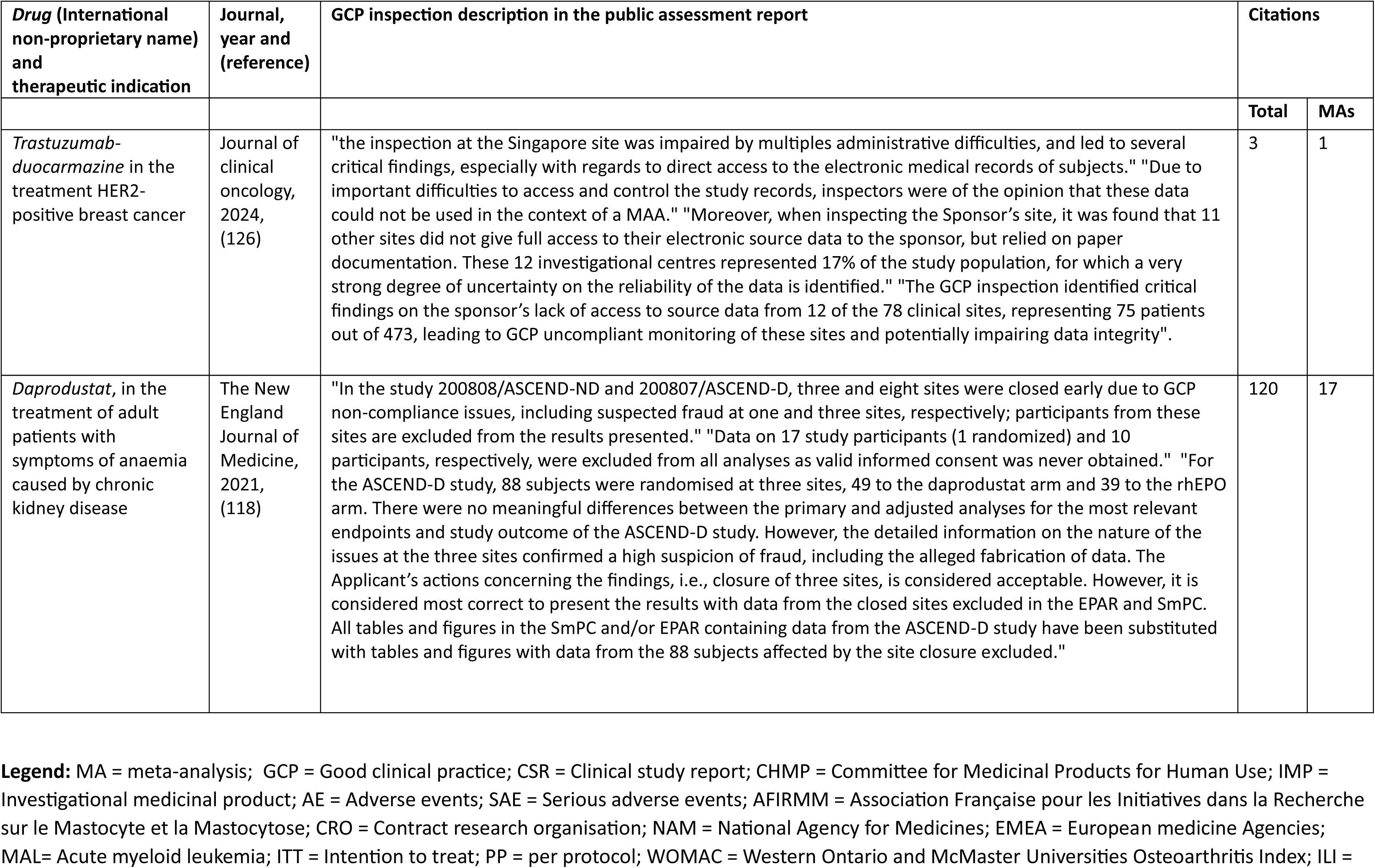
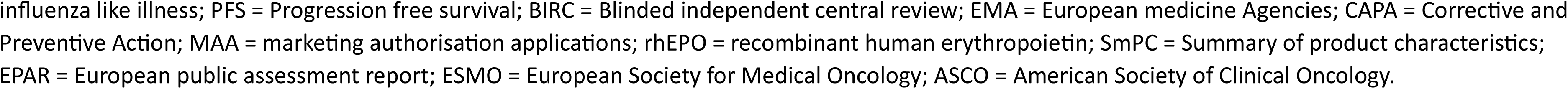
Studies with inspection findings affecting the data reliability. Selected elements of the description of the good clinical practice inspection are provided.

#### Detailed inspection reports

The EMA policy stipulates that “each access to documents request cannot exceed 2 documents”. Our first request for access to GCP inspection reports was made the 4^th^ of December 2024 and concerned emapalumab and daprodustat. To date 2^nd^ of May 2025, we had no answers.

## Discussion

### Statement of principal findings

Among 285 EPARs concerning drugs that were not authorized in the European Union, we found 57 that mentioned a GCP inspection including at least 28 with issues that were considered as affecting reliability of study results. Of those, 24 studies were published in 26 papers. None of those publications acknowledged the issues or were subsequently corrected after publication. While health authorities’ GCP inspection reports serve as a significant source of information, regulators cannot ensure their uptake by the publication ecosystem. This is particularly regrettable given that the medical journals publishing these articles — including the New England Journal of Medicine, The Lancet, and JAMA — are among the most prestigious and are widely regarded as trusted sources of evidence by clinicians, researchers, and public health decision-makers. Many of their readers may be unfamiliar with the highly technical and detailed EPARs and instead rely on journal articles that could fail to provide the full picture for a given drug trial. Journals stand to gain significantly from such independent GCP inspections, which offer a level of scrutiny and depth that far exceeds what peer review alone can deliver. This additional layer of oversight could strengthen the credibility and reliability of the content they publish. We do not believe that all studies with potential reliability issues should be retracted. In some cases, the concerns may affect only a subset of outcomes or analyses (for example for daprodustat (68,118), where exclusion of sites did not change the overall results). Nonetheless, journals have a responsibility to clearly indicate which parts of a study appear robust and which are questionable or irreparably flawed. The list provided in this paper may be an invaluable resource for achieving this objective.

### Strengths and weaknesses of the study

Our study addresses a novel and underexplored topic of high importance when considering the quality of the medical literature and its ability to self-correct. While we adopted a comprehensive approach by examining all rejected or withdrawn EPARs, several limitations should be acknowledged. Despite the EMA’s efforts to promote transparency, EPARs often lack sufficient detail about GCP inspections, leaving the nature and impact of identified issues unclear in many cases.

Due to our conservative data analysis, which only included explicit concerns and excluded “unclear” situations, we may have overlooked some problematic studies. Studies lacking data reliability assessment and GCP findings description should indeed be approached with caution. For example, for vitespen (23,129), the EPAR states that “the inspection concluded that the pivotal phase 3, randomized, open-label, international clinical trial (C-100-12) was conducted in accordance with Good Clinical Practices. During these inspections, no critical findings were observed but 17 major findings were reported, 3 of which impacted on the results.” As there was no mention of data reliability issues, the exact impact on the findings was not precisely described, and the EMA used the data for analysis, we included this study in the “unknown” reliability category. Another example could be biotin for the treatment of progressive multiple sclerosis for multiple sclerosis (56,125), where this seemingly contradictory sentence can be found: “while there are a number of findings that may have impacted the rights of the patients, the quality and integrity of the data and potentially the results of the trial, the source data appeared to be reliable.” As there was a mention that source data are reliable the exact impact on the findings was not precisely described, and the EMA used the data for analysis, we included this study in the “not impacting the reliability of the data” category. In these situations, a review of complete GPC inspection records could be beneficial. However, our requests for these reports have proven largely ineffective due to administrative constraints, with delays so long as to render the process practically unusable.

To facilitate the identification of EPARs that may contain GCP-related issues, we focused our analysis on refused and withdrawn EPARs. This choice has important implications for both internal and external validity. Regarding internal validity, this approach may introduce selection bias and overestimate the frequency or severity of problems. It is reasonable to assume that studies supporting approved drugs are generally more robust, as these drugs are expected to be backed by more trustworthy evidence and less likely to have critical GCP findings impacting the data, even if this remains to be proven.

In terms of external validity, the clinical relevance of our findings may be limited, as these drugs are not approved for use in the European market. However, this limitation is mitigated by the facts that some of these products may be used off-label, some are marketed in other regions (e.g., ceftobiprole (26), in the USA and Canada), all potentially serve as the basis for further research concerning broader pharmacological class (e.g., Naproxinod (28), or treprostinil sodium (24)) and some may be ultimately marketed in European Union (EU) following a new market application (e.g., abaloparatide (57)). Furthermore, our focus was on the reliability of the published studies’ records, which is relevant regardless of drug approval status. It is also important to note that our focus was on drugs undergoing market authorisation submission, thus our analysis concerned mostly large pivotal trials sponsored by pharmaceutical companies. Consequently, our findings may not be applicable to other studies, including those conducted in academic settings, which may also be subject to various audit procedures.

### Strengths and weaknesses in relation to other studies, discussing important differences in results

Bearing these limitations in mind, our study aligns with and replicates findings from the previous analysis of FDA data by Seife (6) with several subtle differences. Both studies found that violations of GCP identified by health authorities are almost never reported in peer-reviewed literature. In addition, these analyses offer complementary insights; Seife’s analysis provides detailed observations, but sometimes lacks clarity on the exact impact of GPC violations on study results (6). As illustrated in our study, health authorities might have identified “critical findings” (EMA) or “official action indicated” (the most severe classification by the FDA (6)) that do not affect the data reliability or that are not clinically relevant. Without direct access to GPC inspections, our analysis is less precise, but CHMP’s assessment of impact on study results helps identify studies critically needing mention of inspections in their publication. Despite challenges in following a few CHMP opinions (e.g., biotin (56) and vitespen (23)), where the EMA’s verbatim text seems to contradict its own conclusions), both GCP reports and expert committee analyses are valuable for end users. Importantly, only one study (28,131) was included in both Seife’s and our analyses, with different yet compatible conclusions. In Seife’s study, the findings of the inspection report were classified as falsification and protocol violations, but no definitive link could be made with the reliability of the published study (6). In our study, the EPAR clearly describe the relation of the findings with the involved study on naproxinod for knee osteoarthritis (131). However, while protocol violations regarding inclusion criteria were acknowledged, falsification was not mentioned in the EPAR.

Our findings are also consistent with a substantial body of literature on reporting bias, which suggests that data from health authorities provide a much more accurate picture than published data—most notably illustrated by the seminal study on publication bias in antidepressant trials where most trials with “negative” results in FDA records were either unpublished or published with “positive” results in the journal literature (133).

### Meaning of the study: possible explanations and implications for clinicians and policymakers

All these results show a clear gap between health authorities’ data and medical literature. First, journal processes appear suboptimal, particularly for correcting the record. There is considerable data on the limitations of post-publication peer review as currently implemented (134–137). While most medical journal editors follow the Committee for Publication Ethics recommendations (138), which advocate for and facilitate post-publication comments and offer guidance on addressing criticisms of articles, such comments are often not sufficiently encouraged by journals. Journals often enforce editorial policies that set word or time constraints following the initial publication of an article (139). Journals occasionally offer online comments but those often lack visibility. We did not systematically search for such comments, as they are beyond the scope of our study but we found two studies (about anamorelin (124) and inactivated influenza virus vaccine for children (128)) that were subject of online comments about the GCP inspection (140,141). These comments explicitly pointed out the issues with data reliability. However, no correction was made to the studies despite those comments. Consequently, post-publication peer review is underdeveloped and often ineffective. For example, insufficient actions were taken when instances of outcome switching were identified and reported to the editors of leading medical journals (135). This highlights the need for change. In our specific context, the added value of GCP inspection reports warrants the implementation of several targeted actions. When the inspection precedes publication of the clinical trial, reporting guidelines such as CONSORT (142) could be more explicit about strongly recommending the report of such inspections. Accordingly, journal policies must require reporting of any GCP findings that questions data reliability. If the inspection occurs after publication, journals should require authors to correct the record in case of any GCP inspection raising concerns about data reliability of their study. COPE could clarify that failing to report and address GCP outcomes questioning study reliability constitutes scientific misconduct as it violates ethical standards. However, it remains to be determined who should be responsible for communicating GCP inspections findings to medical journals. Open science models connecting the various institutions involved in clinical trial governance - including health authorities - with the publication records have been proposed (143,144).

Authors of research papers have a responsibility to report critical CGP irregularities to the editors in a clear and timely manner. However, the case of the RECORD-4 trial (145) published in 2009, has revealed limitations in this approach even in the rare case where a correction was made. Several significant issues were identified by the FDA in 2014 and they were also brought to light by Seife’s investigation in 2015 (6). However, the RECORD-4 publication was not corrected until 2022, when the first author acknowledged that “in October 2022, concerns about the accuracy of data presented in the article were brought to [his] attention” and that the FDA report had been “previously unknown to the RECORD4 Steering Committee” (146).

An alternative approach would be to rely on sponsors to declare information about GCP inspections to journals. Initiatives like Good Pharma scorecard (147) may help in this regard by providing adequate incentives. This project aims to monitor desirable aspects of clinical research among pharmaceutical companies to enhance their practices. Transparency about GCPs inspections may reinforce trust in the company, not only for its ability to correct the record when needed, but also, in a more positive way, when those reports show no concerns. However, relying on sponsors may be seen as wishful thinking, considering the inherent conflicts of interest at play (148).

Therefore, in the absence of concrete mechanisms for accounting for GCP inspections outcomes in the medical literature, we believe that studies like this one and initiatives like PubPeer (a site dedicated to post publication peer review) (11) are essential to raise awareness until systemic action is implemented. Unfortunately, Pubpeer appeared in our study to be underused. To support such initiatives the least health authorities such as EMA can do is to publicly release their inspection findings. However, there is currently no publicly available list at EMA.

### Unanswered questions and future research

This systematic analysis of GCP inspections, as documented in the EPARs of drugs not authorized in the European Union, identified several studies published in the medical literature that warrant correction. These studies likely convey inaccurate messages, even potentially exposing patients to off-label use of drugs without a clear understanding of their harm–benefit ratio or steering the research community toward misguided directions. Future research projects that should analyse full GCP inspection reports—when available—and extend this line of inquiry to drugs approved by the EMA. Further research should also focus on developing effective strategies for correcting the scientific record.

#### What is already known on this topic

In evidence-based medicine, reliable decision making relies on the best available evidence, usually from the medical literature

However, the pre-publication peer review system is known to have several weaknesses, particularly not being able to conduct inspection on trial sites.

Good clinical practice inspections performed by the Food and Drug Administration are unfrequently reported in the medical literature.

#### What this study adds

This study showed that using the data from the European Medicine Agency is a valuable way to identify published studies, that may need corrections.

The data from the European Medicine Agency are complementary to those from the Food and Drug Administration.

The post-publication peer review system still needs improvements, to offer the best evidences for decision making.

## Supporting information

Supplementary references and Supplementary Tables 1, 2 and 3

## Ethics statements

Ethical approval Not required.

