## Supplementary references and Supplementary Tables 1, 2 and 3 for "Concordance between EMA Good Clinical Practice inspections and medical literature concerning drugs that have not received marketing authorization in the European Union: a meta-research survey": 20250509 Supplementary references and Tables 1 2 3.pdf

**Supplementary material**

**Supplementary legend:**

**Supplementary Table 1**: List of EPAR and description of good clinical practice inspection findings.

Legend: * Sponsor clinical number; GVHD = graft versus host disease; ± = EudraCT trial number; ‡ = National Clinical Trial; - = not mentioned; NA= Not available.

**Supplementary Table 2:** EPAR and publications paired, description of the relevance of the good clinical practice inspection findings on the data reliability and diffusion of the studies in the literature.

Legend: * = meeting abstract; GVHD = graft versus host disease; ESMO = European Society for Medical Oncology; ASCO = American Society of Clinical Oncology.

**Supplementary Table 3:** List of unpublished studies and impact of the good clinical practice inspection findings on the data reliability.

Legend: * Sponsor clinical number; ± = EudraCT trial number; ‡ = National Clinical Trial; NA= Not available.

**Supplementary references:**

**Supplementary Table 1**: List of EPAR and description of good clinical practice inspection findings.

| Drug (commercial, used in EPAR) | Drug (International non-proprietary name) | Deviation | | | Concerns about | | | Number of critical deviation | Related studies | Number of articles related | EPAR reference |
|---|---|---|---|---|---|---|---|---|---|---|---|
| | | Critical | Major | Minor | Efficacy | Safety | Ethic | | | | |
| Alpheon | Interferon alfa-2a | - | - | - | - | - | - | NA | BP-IFN-002* | 0 | (17) |
| Mycograb | Efungumab | Yes | Yes | - | Yes | Yes | Yes | NA | NA | 1 | (13) |
| Zeftera | Ceftobiprole | - | - | - | Yes | Yes | - | NA | 2004-001662-41$^{\pm}$ ; 2005-002301-23$^{\pm}$ | 1 | (26) |
| Sumatriptan Galpharm | Sumatriptan | - | - | - | - | - | - | NA | 03-115* | 0 | (30) |
| Istodax | Romidepsin | - | Yes | - | Yes | - | - | 0 | GPI-06-0002* | 1 | (32) |
| Labazenit | budesonide / salmeterol | Yes | - | - | Yes | - | - | NA | BUSAL SS071* ; BUSAL III-02-01* | 0 | (36) |
| Masican | Masitinib | Yes | Yes | - | Yes | Yes | - | 23 | AB07001* | 1 | (39) |
| Reasanz | Serelaxin | - | Yes | - | - | - | - | 0 | NCT00520806$^{‡}$ | 1 | (40) |
| Heparesc | Human heterologous liver cells | Yes | Yes | - | Yes | Yes | - | 11 | NCT00718627$^{‡}$ | 1 | (45) |
| Human IGG1 monoclonal antibody specific for human interleukin-1 alpha XBiotech | Human IgG1 monoclonal antibody specific for human interleukin-1 alpha | - | - | - | - | - | - | NA | 2014-000550-12$^{\pm}$ | 1 | (53) |

| Drug (commercial, used in EPAR) | Drug (International non-proprietary name) | Deviation | | | Concerns about | | | Number of critical deviation | Related studies | Number of articles related | EPAR reference |
|---|---|---|---|---|---|---|---|---|---|---|---|
| | | Critical | Major | Minor | Efficacy | Safety | Ethic | | | | |
| Adlumiz | Anamorelin | - | - | - | Yes | Yes | - | NA | NCT01387269‡ ; NCT01387282‡ | 1 | (52) |
| Masipro | Masitinib | Yes | Yes | Yes | Yes | Yes | - | NA | NCT00814073‡ | 1 | (54) |
| Xeljanz | Tofacitinib | Yes | Yes | - | - | - | - | NA | NCT00814307‡; NCT00814307‡; NCT00847613‡ | 3 | (38) |
| Alsitek | Masitinib | Yes | Yes | Yes | Yes | Yes | - | 4 | NCT02588677‡ | 2 | (58) |
| Eladynos | Abaloparatide | Yes | - | - | - | - | - | NA | NCT01343004‡ | 1 | (57) |
| Gamifant | Emapalumab | Yes | Yes | - | Yes | Yes | Yes | NA | NCT01818492‡ | 1 | (64) |
| Nouryant | Istradefylline | - | - | - | - | - | - | NA | NCT00456586‡; NCT00456794‡; 6002-US-013*; 6002-0608*; NCT00955526‡; 6002-US-018*; NCT00199394‡; NCT01968031‡ | 6 | (66) |
| Arxxant | Ruboxistaurin | - | - | - | - | - | - | NA | NCT00604383‡ | 1 | (14) |
| Cerepro | Herpes simplex virus thymidine kinase gene | - | - | - | - | - | - | NA | Study 903* | 1 | (15) |
| Retisert | Fluocinolone acetonide | - | - | - | - | - | - | NA | BLP 415-002* | 1 | (16) |
| Kiacta | Eprodisate disodium | - | - | - | Yes | - | - | NA | NCT00035334‡ | 1 | (18) |

| **Drug (commercial, used in EPAR)** | **Drug (International non-proprietary name)** | **Deviation** | | | **Concerns about** | | | **Number of critical deviation** | **Related studies** | **Numbe r of articles related** | **EPAR reference** |
|---|---|---|---|---|---|---|---|---|---|---|---|
| | | **Critical** | **Major** | **Minor** | **Efficacy** | **Safety** | **Ethic** | | | | |
| Lenalidomide Celgene Europe | Lenalidomide | - | - | - | Yes | Yes | - | NA | NCT00065156[‡] | 1 | (20) |
| Aflunov | A/VietNam/1194/2004 vaccine | - | - | - | Yes | Yes | Yes | NA | 2006-005428-18[±] | 0 | (19) |
| Vekacia | Ciclosporin | - | - | - | - | - | - | NA | NCT00328653[‡] | 1 | (21) |
| Nenad | Lisuride | - | - | - | - | - | - | NA | TULIP-IIb* | 0 | (22) |
| Oncophage | Vitespen | - | Yes | - | - | - | - | 0 | NCT00033904[‡] | 1 | (23) |
| Repaglinide Sun | Repaglinide | - | Yes | - | - | - | - | 0 | PKD_08_059* | 0 | (25) |
| Tyvaso | Treprostinil sodium | Yes | Yes | - | Yes | Yes | - | 6 | NCT00147199[‡] | 1 | (24) |
| Zenhale | Mometasone furoate and formoterol fumarate | Yes | Yes | - | - | - | - | NA | NCT00383240[‡] | 1 | (27) |
| Beprana | Naproxcinod | Yes | - | - | Yes | - | - | NA | NCT00542555[‡]; NCT00504127[‡]; NCT00541489[‡] | 3 | (28) |
| Luveniq | Voclosporin | Yes | Yes | - | Yes | Yes | - | NA | 2006-006543-31[±]; 2006-006544-66[±] | 1 | (29) |
| Fluad Paediatric | Inactivated influenza (flu) virus strains | Yes | Yes | - | Yes | Yes | - | 3 | NCT00644059[‡] | 1 | (31) |
| Loulla | Mercaptopurine | - | - | - | - | - | - | NA | NA | 0 | (34) |
| Jenzyl | Ridaforolimus | - | Yes | - | Yes | - | - | 0 | NCT00538239[‡] | 1 | (33) |

| Drug (commercial, used in EPAR) | Drug (International non-proprietary name) | Deviation | | | Concerns about | | | Number of critical deviation | Related studies | Number of articles related | EPAR reference |
|---|---|---|---|---|---|---|---|---|---|---|---|
| | | Critical | Major | Minor | Efficacy | Safety | Ethic | | | | |
| Omontys | Peginesatide | - | - | - | Yes | - | - | NA | NCT00597753[‡]; NCT00597584[‡] | 1 | (37) |
| OraNera | Autologous oral mucosal epithelial cells | Yes | Yes | - | - | Yes | - | NA | 2007-A00270-53[±] | 1 | (35) |
| Heplisav | Vaccine which contains hepatitis B surface antigen | - | - | - | Yes | Yes | - | NA | NCT00985426[‡] | 1 | (41) |
| Neocepri | Folic acid | - | - | - | - | - | - | NA | NA | 0 | (44) |
| Vynfinit | Vintafolide | - | - | - | - | - | - | NA | NCT00722592[‡] | 1 | (43) |
| Folcepri | Etarfolatide | - | - | - | - | - | - | NA | NA | 0 | (42) |
| Veraseal | Human fibrinogen and human thrombin | Yes | - | - | Yes | Yes | - | NA | NCT01754480[‡] | 1 | (46) |
| Kyndrisa | Drisapersen | - | - | - | - | Yes | - | NA | NCT01480245[‡] | 0 | (48) |
| Opsiria | Sirolimus | - | - | - | - | - | - | 0 | NCT01358266[‡] | 1 | (47) |
| Zemfirza | Cediranib | - | - | - | Yes | Yes | - | NA | NCT00532194[‡] | 1 | (49) |
| Graspa | Eryaspase | - | Yes | - | - | - | Yes | 0 | NCT01518517[‡] | 1 | (51) |
| Efgratin | Pegfilgrastim | - | Yes | Yes | - | - | - | 0 | 2013-003166-14[±] | 1 | (50) |
| Qinprezo | Vosaroxin | - | Yes | Yes | - | - | - | 0 | NCT01191801[‡] | 1 | (55) |
| Qizenday | Biotin | - | Yes | - | Yes | - | - | 0 | 2013-002113-35[±] | 1 | (56) |
| Nuzyra | Omadacycline | - | - | - | - | - | - | NA | NCT02378480[‡]; NCT02531438[‡] | 2 | (59) |
| Idhifa | Enasidenib | Yes | Yes | - | - | - | - | NA | NCT01915498[‡] | 2 | (61) |

| **Drug (commercial, used in EPAR)** | **Drug (International non-proprietary name)** | **Deviation** | | | **Concerns about** | | | **Number of critical deviation** | **Related studies** | **Numbe r of articles related** | **EPAR reference** |
|---|---|---|---|---|---|---|---|---|---|---|---|
| | | **Critical** | **Major** | **Minor** | **Efficacy** | **Safety** | **Ethic** | | | | |
| Luxceptar | T-cells from the transplant donor, specially treated to reduce the risk of GVHD | - | Yes | - | - | - | - | 0 | NCT01794299[‡] | 2 | (60) |
| Doxorubicin Hydrochloride Tillomed | Doxorubicin | Yes | Yes | - | Yes | - | - | 2 | PLCL 200 17* | 0 | (62) |
| Sondelbay | Teriparatide | - | - | - | - | - | - | NA | 0425-17* | 0 | (63) |
| Upkanz | Deferiprone | - | Yes | Yes | - | - | - | 0 | NCT01741532[‡] | 1 | (65) |
| Tuznue | Trastuzumab | Yes | Yes | - | Yes | Yes | - | NA | 2016-004019-11[±] | 2 | (67) |
| Jivadco | Trastuzumab-duocarmazine | Yes | Yes | - | Yes | Yes | - | NA | NCT03262935[‡] | 2 | (69) |
| Jesduvroq | Daprodustat | - | - | - | - | - | - | NA | NCT02876835[‡]; NCT02879305[‡] | 2 | (68) |

Table Legend : GVHD = graft versus host disease; * Sponsor clinical number; ± = EudraCT trial number; ‡ = National Clinical Trial; - = not mentioned; NA= Not available.

**Supplementary Table 2:** EPAR and publications paired, description of the relevance of the good clinical practice inspection findings on the data reliability and diffusion of the studies in the literature.

| Drug (commercial) (EPAR reference) | Drug (International non-proprietary name) (publication reference) | PMID | DOI | Journal | Number of citations | Citation by Meta-analysis | Data Reliability |
|---|---|---|---|---|---|---|---|
| Mycograb (13) | Efungumab (108) | 16619152 | 10.1086/503428 | Clinical Infectious Diseases | 227 | 2 | Not affected |
| Zeftera (26) | Ceftobiprole (106) | 18225981 | 10.1086/526527 | Clinical Infectious Diseases | 208 | 12 | Affected |
| Istodax (32) | Romidepsin (75) | 22271479 | 10.1200/JCO.2011.37.4223 | Journal of clinical oncology | 533 | 6 | Unknown |
| Masican (39) | Masitinib (70) | 25122671 | 10.1093/annonc/mdu237 | Annals of oncology | 54 | 6 | Affected |
| Reasanz (40) | Serelaxin (123) | 23141816 | 10.1016/S0140-6736(12)61855-8 | The Lancet | 702 | 3 | Not affected |
| Heparesc (45) | human heterologous liver cells (97) | 29027067 | 10.1007/s10545-017-0097-4 | Journal of Inherited Metabolic Disease | 14 | 0 | Affected |
| Human IGG1 monoclonal antibody specific for human interleukin-1 alpha XBiotech (53) | human IgG1 monoclonal antibody specific for human interleukin-1 alpha (83) | 28094194 | 10.1016/S1470-2045(17)30006-2 | The Lancet Oncology | 120 | 1 | Unknown |
| Adlumiz (52) | Anamorelin (124) | 26906526 | 10.1016/S1470-2045(15)00558-6 | The Lancet Oncology | 415 | 9 | Affected |
| Masipro (54) | Masitinib (94) | 28069279 | 10.1016/S0140-6736(16)31403-9 | The Lancet | 93 | 2 | Affected |
| Xeljanz (38) | Tofacitinib (81) | 22873530 | 10.1056/NEJMoa1109071 | The New England Journal of Medicine | 742 | 44 | Not affected |
| Xeljanz (38) | Tofacitinib (122) | 26530039 | 10.1186/s13075-015-0825-9 | Arthritis Research & Therapy | 51 | 4 | Not affected |
| Xeljanz (38) | Tofacitinib (127) | 23348607 | 10.1002/art.37816 | Arthritis & Rheumatism | 455 | 42 | Affected |

| Drug (commercial) (EPAR reference) | Drug (International non-proprietary name) (publication reference) | PMID | DOI | Journal | Number of citations | Citation by Meta-analysis | Data Reliability |
|---|---|---|---|---|---|---|---|
| Alsitek (58) | Masitinib (102) | 31280619 | 10.1080/21678421.2019.1632346 | Amyotrophic Lateral Sclerosis and Frontotemporal Degeneration | 135 | 1 | Affected |
| Alsitek (58) | Masitinib (101) | 34457038 | 10.1177/17562864211030365 | Therapeutic Advances in Neurological Disorders | 38 | 0 | Affected |
| Eladynos (57) | Abaloparatide (98) | 27533157 | 10.1001/jama.2016.11136 | JAMA | 571 | 25 | Affected |
| Gamifant (64) | Emapalumab (93) | 32374962 | 10.1056/NEJMoa1911326 | The New England Journal of Medicine | 319 | 0 | Affected |
| Nouryant (66) | Istradefylline (91) | 18306243 | 10.1002/ana.21315 | Annals of Neurology | 284 | 8 | Unknown |
| Nouryant (66) | Istradefylline (119) | 18519872 | 10.1212/01.wnl.0000313834.22171.17 | Neurology | 145 | 6 | Unknown |
| Nouryant (66) | Istradefylline (82) | 18831530 | 10.1002/mds.22095 | Movement Disorders | 167 | 8 | Unknown |
| Nouryant (66) | Istradefylline (99) | 20629136 | 10.1002/mds.23107 | Movement Disorders | 141 | 8 | Unknown |
| Nouryant (66) | Istradefylline (100) | 23483627 | 10.1002/mds.25418 | Movement Disorders | 167 | 6 | Unknown |
| Nouryant (66) | Istradefylline (110) | 22000279 | 10.1016/j.parkreldis.2011.09.023 | Parkinsonism Related disoders | 88 | 8 | Unknown |
| Arxxant (14) | Ruboxistaurin (76) | 18708615 | 10.1167/iovs.08-2473 | Investigative Ophthalmology & Visual Science | 64 | 0 | Unknown |
| Cerepro (15) | Herpes simplex virus thymidine kinase gene (85) | 15509514 | 10.1016/j.ymthe.2004.08.002 | Molecular Therapy | 298 | 5 | Affected |
| Retisert (16) | Fluocinolone acetonide (109) | 20079922 | 10.1016/j.ophtha.2009.11.027 | Ophtalmology | 110 | 2 | Unknown |
| Kiacta (18) | Eprodisate disodium (78) | 17554116 | 10.1056/NEJMoa065644 | The New England Journal of Medicine | 162 | 1 | Affected |

| Drug (commercial) (EPAR reference) | Drug (International non-proprietary name) (publication reference) | PMID | DOI | Journal | Number of citations | Citation by Meta-analysis | Data Reliability |
|---|---|---|---|---|---|---|---|
| Lenalidomide Celgene Europe (20) | Lenalidomide (92) | 17021321 | 10.1056/NEJMoa061292 | The New England Journal of Medicine | 1009 | 6 | Affected |
| Vekacia (21) | Ciclosporin (90) | 37872059 | 10.1016/j.clinthera.2023.09.022 | Clinical Therapeutics | 2 | 0 | Unknown |
| Oncophage (23) | Vitespen | 18602688 (129) | 10.1016/S0140-6736(08)60697-2 | The Lancet | 263 | 6 | Unknown |
| Tyvaso (24) | Treprostinil sodium (96) | 20430262 | 10.1016/j.jacc.2010.01.027 | Journal of the American College of Cardiology | 422 | 30 | Affected |
| Zenhale (27) | Mometasone furoate and formoterol fumarate (103) | 20678306 | 10.2500/aap.2010.31.3364 | Allergy and Asthma Proceedings | 26 | 5 | Affected |
| Beprana (28) | Naproxcinod (115) | 20202489 | 10.1016/j.joca.2009.12.013 | Osteoarthritis and Cartilage | 38 | 5 | Affected |
| Beprana (28) | Naproxcinod (116) | 20828790 | 10.1016/j.semarthrit.2010.06.002 | Seminars in Arthritis and Rheumatism | 33 | 4 | Unknown |
| Beprana (28) | Naproxcinod (71) | 20722026 | 10.1002/art.27694 | Arthritis & Rheumatology | 28 | 3 | Not affected |
| Luveniq (29) | Voclosporin (112) | NA | NA (Investigative Ophthalmology & Visual Science April 2009, Vol.50, issue 13) | Investigative Ophthalmology & Visual Science* | NA | NA | Unknown |
| Fluad Paediatric (31) | Inactivated influenza (flu) virus strains (128) | 21995388 | 10.1056/NEJMoa1010331 | The New England Journal of Medicine | 244 | 10 | Affected |
| Jenzyl (33) | Ridaforolimus (79) | 23715582 | 10.1200/JCO.2012.45.5766 | Journal of Clinical Oncology | 179 | 10 | Not affected |
| Omontys (37) | Peginesatide (80) | 23343061 | 10.1056/NEJMoa1203165 | The New England Journal of Medicine | 71 | 2 | Affected |

| Drug (commercial) (EPAR reference) | Drug (International non-proprietary name) (publication reference) | PMID | DOI | Journal | Number of citations | Citation by Meta-analysis | Data Reliability |
|---|---|---|---|---|---|---|---|
| OraNera (35) | Autologous oral mucosal epithelial cells (73) | 22064987 | 10.1167/iovs.11-7744 | Investigative ophtalmology & visual science | 133 | 0 | Not affected |
| Heplisav (41) | vaccine which contains hepatitis B surface antigen (86) | 23727422 | 10.1016/j.vaccine.2013.05.067 | Vaccine | 56 | 1 | Affected |
| Vynfinit (43) | Vintafolide (104) | 24127448 | 10.1200/JCO.2013.49.7685 | Journal of Clinical Oncology | 151 | 4 | Not affected |
| Veraseal (46) | Human fibrinogen and human thrombin (74) | 28647631 | 10.1016/j.avsg.2017.06.043 | Annals of Vascular Surgery | 17 | 4 | Affected |
| Opsiria (47) | Sirolimus (105) | 27692526 | 10.1016/j.ophtha.2016.07.029 | Ophthalmology | 60 | 0 | Not affected |
| Zemfirza (49) | Cediranib (89) | 27025186 | 10.1016/S0140-6736(15)01167-8 | The Lancet | 193 | 15 | Affected |
| Graspa (51) | Eryaspase (72) | NA | 10.1182/blood.V126.23.3723.3723 | ASH (Volume 126, Issue 23, 3 December 2015, Page 3723 ; Blood)* | NA | NA | Not affected |
| Efgratin (50) | Pegfilgrastim (87) | 30727980 | 10.1186/s12885-019-5329-6 | BMC Cancer | 5 | 1 | Not affected |
| Qinprezo (55) | Vosaroxin (111) | 26234174 | 10.1016/S1470-2045(15)00201-6 | Lancet Oncology | 124 | 2 | Not affected |
| Qizenday (56) | Biotin (125) | 27589059 | 10.1177/1352458516667568 | Multiple Sclerosis Journal | 221 | 0 | Not affected |
| Nuzyra (59) | Omadacycline (107) | 30726689 | 10.1056/NEJMoa1800170 | The New England Journal of Medicine | 119 | 7 | Not affected |
| Nuzyra (59) | Omadacycline (121) | 30726692 | 10.1056/NEJMoa1800201 | The New England Journal of Medicine | 155 | 4 | Not affected |
| Idhifa (61) | Enasidenib (120) | 28588020 | 10.1182/blood-2017-04-779405 | Blood | 1095 | 3 | Not affected |

| Drug (commercial) (EPAR reference) | Drug (International non-proprietary name) (publication reference) | PMID | DOI | Journal | Number of citations | Citation by Meta-analysis | Data Reliability |
|---|---|---|---|---|---|---|---|
| Idhifa (61) | Enasidenib (77) | 34427990 | 10.1002/cam4.4182 | Cancer med | 6 | 0 | Not affected |
| Luxceptar (60) | T-cells from the transplant donor, specially treated to reduce the risk of GVHD (114) | NA | 10.1182/blood-2018-99-119086 | Blood | NA | NA | Not affected |
| Luxceptar (60) | T-cells from the transplant donor, specially treated to reduce the risk of GVHD (114) | 32047237 | 10.1038/s41375-020-0733-0 | Leukemia | 19 | 0 | Not affected |
| Upkanz (65) | Deferiprone (88) | 31202468 | 10.1016/S1474-4422(19)30142-5 | Lancet Neurology | 90 | 1 | Not affected |
| Tuznue (67) | Trastuzumab (130) | NA | 10.1016/j.annonc.2020.08.288 | Annals of oncology (ESMO abstract)* | NA | NA | Affected |
| Tuznue (67) | Trastuzumab (84) | NA | 10.1200/JCO.2020.38.15_suppl.579 | Journal of clinical oncology (ASCO abstract)* | NA | NA | Affected |
| Jivadco (69) | Trastuzumab-duocarmazine (95) | NA | 10.1016/j.annonc.2021.08.2088 | Annals of Oncology (ESMO abstract)* | NA | NA | Affected |
| Jivadco (69) | trastuzumab-duocarmazine (126) | 39442070 | 10.1200/JCO.24.00529 | Journal of clinical oncology | 3 | 1 | Affected |
| Jesduvroq (68) | Daprodustat (117) | 34739196 | 10.1056/NEJMoa2113380 | The New England Journal of Medicine | 138 | 15 | Not affected |
| Jesduvroq (68) | Daprodustat (118) | 34739194 | 10.1056/NEJMoa2113379 | The New England Journal of Medicine | 120 | 17 | Affected |

Legend: * = meeting abstract; GVHD = graft versus host disease; ESMO = European Society for Medical Oncology; ASCO = American Society of Clinical Oncology.

**Supplementary Table 3:** List of unpublished studies and impact of the good clinical practice inspection findings on the data reliability.

| Drug (commercial) | Drug (International non-proprietary name) | EPAR reference | Related study | Data Reliability |
|---|---|---|---|---|
| Alpheon | interferon alfa-2a | (17) | BP-IFN-002* | Not affected |
| Zeftera | ceftobiprole | (26) | 2004-001662-41± | Affected |
| Sumatriptan Galpharm | sumatriptan | (30) | 03-115* | Affected |
| Labazenit | budesonide / salmeterol | (36) | BUSAL SS071* | Not affected |
| Labazenit | budesonide / salmeterol | (36) | BUSAL III-02-01* | Unknown |
| Nouryant | istradefylline | (66) | NCT00199394‡ | Unknown |
| Nouryant | istradefylline | (66) | NCT01968031‡ | Unknown |
| Aflunov | A/VietNam/1194/2004 vaccine | (19) | 2006-005428-18± | Affected |
| Nenad | lisuride | (22) | TULIP-IIb* | Unknown |
| Repaglinide Sun | repaglinide | (25) | PKD_08_059* | Unknown |
| Loulla | mercaptopurine | (34) | NA | Affected |
| Neocepri | folic acid | (44) | NA | Not affected |
| Folcepri | etarfolatide | (42) | NA | Not affected |
| Kyndrisa | drisapersen | (48) | NCT01480245‡ | Not affected |
| Doxorubicin Hydrochloride Tillomed | doxorubicin | (62) | PLCL 200 17* | Affected |
| Sondelbay | teriparatide | (63) | 0425-17* | Affected |

Legend: * Sponsor clinical number; ± = EudraCT trial number; ‡ = National Clinical Trial; NA= Not available.
